# Patient and Clinician Perspectives on Centralized Cascade Screening for Familial Hypercholesterolemia in the United States: A Qualitative Implementation Study

**DOI:** 10.64898/2026.08.18.26359725

**Authors:** Megan C. Roberts, Laney K. Jones, Ashley Brown, Jessica Carda-Auten, Marina Cuchel, Alison R. Hilton, Amit Khera, Mark Rothstein, Kyaw Soe, Aurora Sullivan, Eric P. Tricou, Maihan B. Vu, William Weintraub, Zahid Ahmad

**Author notes:** **Corresponding author**: Zahid Ahmad, MD.

## Abstract

**Objective:** To identify patient- and clinician-reported barriers, facilitators, and design requirements for a centralized cascade-screening program for familial hypercholesterolemia (FH) in the United States.

**Methods:** From June through November 2023, we conducted individual telephone interviews with 20 patients with FH and 10 clinicians recruited from UT Southwestern Medical Center, Parkland Health, the North Texas Veterans Affairs, and other clinical settings. Interview guides were informed by the Consolidated Framework for Implementation Research. Transcripts were coded in Dedoose using a piloted codebook, with discrepancies and emergent themes resolved through consensus. An advisory panel then helped translate interview findings into program design requirements and implementation strategies.

**Results:** Five themes characterized barriers and facilitators to centralized cascade screening: (1) health-system access and fragmentation, including screening and treatment costs, transportation, and cross-system coordination; (2) privacy and trust, including concerns about genetic information and unsolicited outreach; (3) family relationships and practical burden, including competing demands, language barriers, limited contact, fear, and denial; (4) clinician capacity and workflow, including limited time, knowledge, and genetic-counseling capacity; and (5) communication and care continuity. Participants recommended proband pre-notification of relatives, culturally and linguistically responsive materials, secure data exchange, standardized scripts, flexible testing pathways, and centralized coordination. These findings informed a program model incorporating a secure pedigree platform, educational and communication resources, testing coordination, and linkage to follow-up care.

**Conclusions:** Patients and clinicians identified multilevel determinants that a centralized FH cascade-screening program must address. The findings support specific design requirements but do not establish program feasibility or effectiveness, which require prospective evaluation.

**Policy implications:** Scaling centralized cascade screening will require attention to affordability, privacy, clinician workflow, culturally responsive communication, and continuity of care across health systems.

## Introduction

Familial hypercholesterolemia (FH) is a common inherited disorder characterized by lifelong elevation of low-density lipoprotein cholesterol and increased risk of premature atherosclerotic cardiovascular disease. Heterozygous FH affects approximately 1 in 250 people, corresponding to an estimated 1.3 million individuals in the United States, yet most remain undiagnosed.^1–3^ Because FH is inherited in an autosomal dominant pattern, each first-degree relative of an affected individual has a 50% probability of also having FH. Cascade screening—the systematic evaluation of at-risk relatives—can identify affected family members early enough to initiate preventive treatment. Public health institutions and professional societies therefore recommend cascade screening as a central strategy for FH detection.^4–10^

In the United States, however, cascade screening generally depends on clinicians counseling patients to notify their own relatives, without a systematic mechanism to ensure that relatives are reached, tested, and connected to care. Centralized programs in other countries have used dedicated coordinating organizations to collect pedigrees, contact families, arrange testing, and return results. A systematic review found that active cascade-testing approaches can identify new FH cases, although implementation models vary substantially.^11^

Recent U.S. initiatives have evaluated family-communication tools, automated messaging, and chatbot-supported engagement.^12,13^ Implementation frameworks such as the Consolidated Framework for Implementation Research (CFIR) can help identify the multilevel determinants that shape adoption.^14^ Other U.S. studies have examined hybrid implementation strategies and pragmatic cascade-testing models.^15,16^ Nevertheless, less is known about how patients and clinicians perceive a centralized model in which an organization outside the usual clinical encounter coordinates outreach, testing, and follow-up across health systems.

We therefore conducted a qualitative study to identify barriers, facilitators, and design requirements for centralized FH cascade screening in the United States. We then used implementation mapping and advisory-panel input to translate these findings into a proposed program model for prospective evaluation.

## Methods

### Study design and setting

We conducted an interview-based qualitative study from June through November 2023 as the needs-assessment component of an implementation-mapping process.^17^ The study focused on a proposed centralized cascade-screening model in which clinicians identify probands with FH and a coordinating organization supports family-history collection, relative notification, testing, return of results, and linkage to care.

### Participants and recruitment

Using convenience sampling, we recruited 20 patients diagnosed with FH from UT Southwestern Medical Center (n=8), Parkland Health (n=3), and the North Texas Veterans Affairs (n=9). These sites were selected because they were intended to participate in a future pilot of the centralized program. We also recruited 10 clinicians from the pilot sites and other clinical settings. Clinician recruitment included purposive identification of genetic counselors and lipid specialists who routinely care for patients with FH, as well as clinicians in primary care and cardiology, to capture perspectives relevant to broader implementation.

### Data collection

Trained qualitative researchers with experience interviewing patients and clinicians conducted individual telephone interviews lasting 45–60 minutes. Separate interview guides for patients and clinicians were informed by CFIR and addressed determinants at the health-system, clinician, patient, and family levels.^14^ Interviews examined anticipated barriers to participating in centralized cascade screening, resources needed to support participation, and recommendations for program design. Participants were offered a $50 gift card after completing the interview. Interviews were professionally transcribed.

### Qualitative analysis

Transcripts were analyzed in Dedoose (SocioCultural Research Consultants, Los Angeles, California). The initial codebook was piloted and refined until coding was replicable across coders. The final codebook was applied to all transcripts. Emergent themes and coding discrepancies were documented and reconciled through discussion and consensus. The team developed code summaries and narrative summaries and compared patient and clinician perspectives to identify convergent and distinct determinants.

### Translation of findings into program design

An advisory panel guided translation of the interview findings into implementation objectives and strategies. The panel included an FH research scientist, a cardiovascular epidemiologist with clinical and research expertise in FH, two implementation-strategy experts, two patients with FH, a legal expert in genetic discrimination, and representatives of a U.S. health insurer. During one-hour virtual meetings, the panel reviewed interview findings, prioritized reach and feasibility as outcomes for a future pilot, identified necessary performance objectives, and refined proposed program materials and workflows. The resulting blueprint was therefore an application of the qualitative findings rather than evidence of program effectiveness.

### Ethical oversight

The study was approved by the institutional review board of UT Southwestern Medical Center (Dallas, TX).

## Results

### Participants and overview

We interviewed 20 patients and 10 clinicians. Participant characteristics are summarized in Table 1. The analysis identified five interrelated themes: health-system access and fragmentation; privacy and trust; family relationships and practical burden; clinician capacity and workflow; and communication and care continuity. Table 2 summarizes the interview-derived barriers, facilitators, and corresponding program design requirements.

**Table 1.**
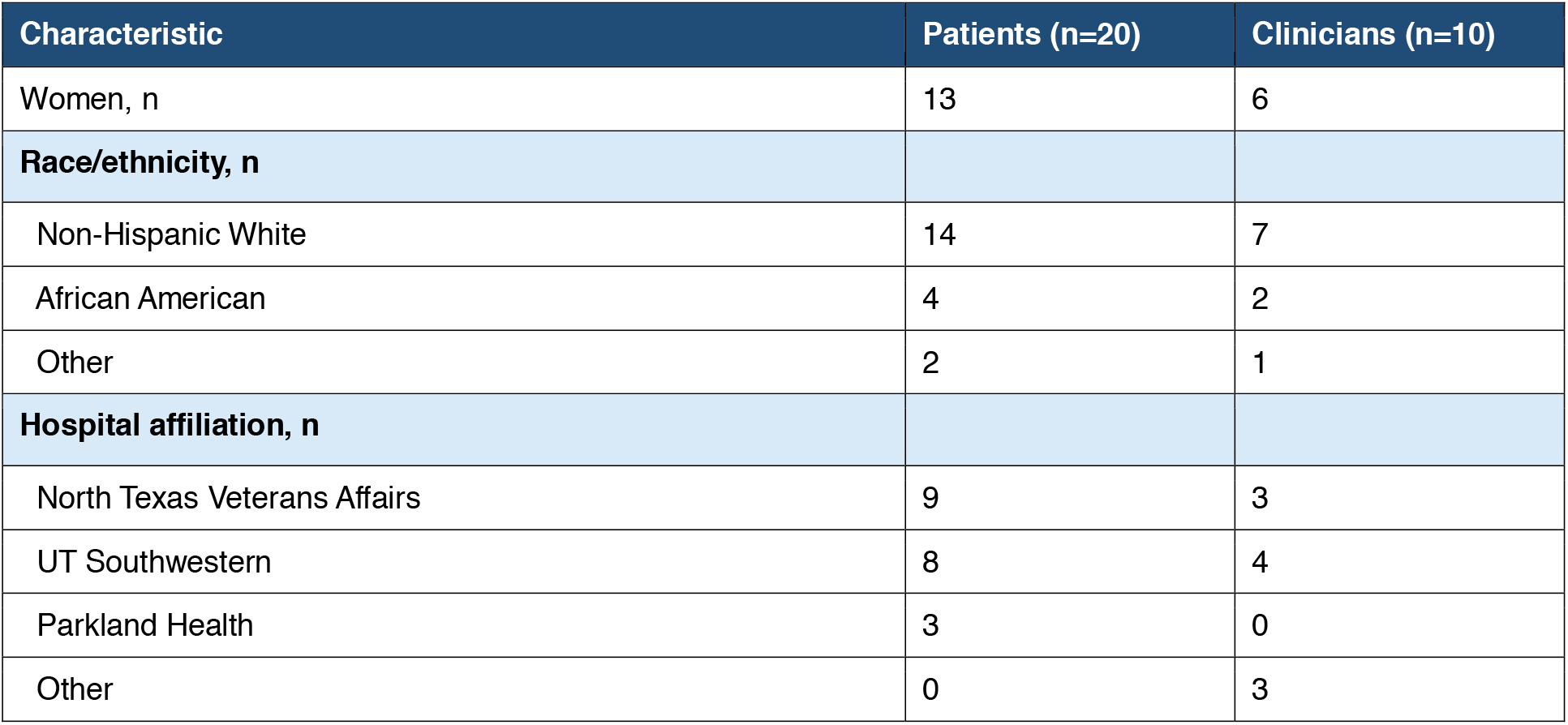
Participant characteristics.

**Table 2.** Interview-derived barriers, facilitators, and program design requirements.

| Theme | Interview findings | Design requirements identified from participant and advisory-panel input |
| --- | --- | --- |
| Health-system access and fragmentation | Screening and treatment costs; geographic distance and transportation; variable access across health systems and for relatives outside the United States. | Transparent information about costs; assistance locating testing and treatment; flexible local or remote pathways; linkage to clinicians and social support after a positive result. |
| Privacy and trust | Uncertainty about permissible contact under HIPAA; concern about genetic-data use; distrust of unfamiliar organizations and unsolicited calls. | HIPAA-compliant data exchange; clear privacy information; recognizable program identity; proband authorization and advance notification before centralized outreach. |
| Family relationships and practical burden | Competing work and caregiving demands; language and communication barriers; limited contact with relatives; denial, fear, or low perceived relevance of FH. | Language-concordant and culturally responsive materials; flexible communication channels; concise education; scripts that help probands notify relatives. |
| Clinician capacity and workflow | Limited visit time; competing clinical priorities; variable knowledge and confidence regarding cascade screening and genetic testing; limited genetic-counseling capacity. | Simple referral processes; standardized scripts and training; automated communication; centralized coordination of counseling, testing, and follow-up where feasible. |
| Communication and care continuity | Patients and clinicians wanted consistent information about FH, the reason for family screening, the screening process, privacy, costs, and next steps after testing. | Shareable patient and family education; provider-facing materials; structured pretest and return-of-results scripts; pathways to preventive care for relatives with positive results. |

#### Theme 1: Health-system access and fragmentation

Patients described screening and treatment costs as linked barriers: identifying FH would have limited value if an affected relative could not subsequently obtain affordable care. As one patient explained, “Even if they are screened that they are positive, being able to get connected with treatment or affordable treatment is another issue as well. Cause not all insurances cover everything.” Distance to testing and follow-up care, transportation, and relatives living outside the United States were additional concerns.

Clinicians similarly anticipated that differences among health systems would complicate both screening and follow-up. Relatives may receive care from organizations that do not share records or use the same testing pathways. Participants therefore viewed coordination as extending beyond test ordering: a centralized program would also need to help relatives identify accessible testing sites and establish appropriate follow-up care after a positive result.

#### Theme 2: Privacy and trust

Clinicians were uncertain about the limits of permissible contact with relatives under the Health Insurance Portability and Accountability Act (HIPAA). One clinician stated, “I mean, our concerns are always with what can we do that is not a HIPAA violation? I think that’s always prevented us from some things.” They emphasized that a centralized model would require clear consent processes, defined responsibilities, and secure information exchange between clinicians and the coordinating organization.

Patients also expressed concern about genetic privacy and the potential use of data outside the clinical relationship. These concerns intersected with broader mistrust of unfamiliar organizations. Participants anticipated that relatives might interpret unsolicited telephone calls, emails, or requests for personal information as fraudulent. One patient explained, “My only thing is you know how people worry about scams these days…First thing is, they’re gonna think it’s a scam.” Advance notification by the proband, recognizable program branding, and transparent explanations of data use were therefore viewed as important facilitators.

#### Theme 3: Family relationships and practical burden

Patients and clinicians described language barriers, cognitive limitations, and competing work and caregiving demands that could impede participation. One patient observed, “They feel like they’re getting pulled and dragged through life as it is, so to add on—especially if they have kids or work trying to make ends meet. It’s just a very inconvenient thing. To take a day off work, that sounds like a huge, huge thing.” Flexible communication and testing pathways were considered important for reducing this burden.

Family relationships also shaped whether risk information could reach relatives. Patients described limited contact with extended family, strained relationships, uncertainty about relatives’ names or contact information, and reluctance to discuss health information. Some anticipated that relatives would minimize the relevance of FH, avoid health care, fear a diagnosis, or respond with denial. These findings suggested that a single outreach pathway would be insufficient and that probands should be able to choose whether they or the coordinating organization initiated contact with each relative.

Participants also recognized that probands may lack the knowledge or confidence to explain FH and cascade screening. They recommended concise scripts, sample emails, and shareable educational materials that could help probands alert relatives before centralized outreach without requiring them to provide genetic counseling themselves.

#### Theme 4: Clinician capacity and workflow

Clinicians described variable knowledge of cascade screening and genetic testing, limited comfort providing genetic counseling, and concern about the scarcity of genetic counselors. One clinician noted that implementing cascade screening outside genetic-counseling settings would be challenging but necessary because “there’s a limited number of genetic counselors.” Time was an equally prominent barrier. Clinicians anticipated difficulty adding family screening discussions to already crowded visits and responding to subsequent electronic health record messages. Competing clinical needs were often perceived as more immediate or more closely aligned with the purpose of a lipid-clinic visit. Participants therefore favored a model that would “keep it simple” for referring clinicians through brief scripts, streamlined referral, secure data transfer, automated communication, and centralized coordination of tasks that did not require the treating clinician.

#### Theme 5: Communication and care continuity

Across participant groups, clear and consistent communication was the most frequently proposed facilitator. Clinicians requested concise education and communication guides addressing genetic testing, cascade screening, and their role in the program. Patients requested materials that could be shared with relatives and that explained FH, the reason for screening, testing options, treatment, the coordinating organization, genetic-privacy protections, potential costs, and how to locate an FH clinician.

Participants emphasized that information should be available in multiple languages and formats, including print, websites, and video, and should be usable by people with different levels of health literacy and access to technology. They also emphasized that communication should continue after testing. Relatives with positive results would need understandable result reports, assistance identifying an appropriate clinician, and access to treatment and social support.

### Translation of qualitative findings into program components

The study team and advisory panel translated these findings into six design requirements: minimize clinician burden; prepare families for centralized outreach; protect privacy and explain data use; address language, knowledge, and trust barriers; reduce testing and navigation barriers; and ensure follow-through after testing. These requirements informed a secure pedigree platform, standardized scripts, educational materials, flexible family-contact pathways, centralized testing coordination, and structured return-of-results and referral processes (Table 3).

**Table 3.** Translation of qualitative findings into components of the centralized cascade-screening model.

| Interview-derived need | Program component | Intended function |
| --- | --- | --- |
| Minimize clinician burden | Brief provider scripts, streamlined referral, and secure electronic transfer to the coordinating center | Limit additional work during time-constrained visits and clarify clinician responsibilities. |
| Prepare families for centralized outreach | Proband primer, sample email or script, and option for proband-led notification | Reduce concern that outreach is a scam and respect family communication preferences. |
| Protect privacy and explain data use | Consent-based, HIPAA-compliant pedigree platform and standardized privacy information | Clarify permissible information sharing and support secure coordination across organizations. |
| Address language, knowledge, and trust barriers | Shareable educational materials for probands, relatives, and clinicians | Provide consistent, culturally responsive information about FH, testing, treatment, costs, and genetic privacy. |
| Reduce logistical barriers | Central assistance locating testing sites and coordinating genetic and lipid testing | Decrease travel and navigation burden and accommodate relatives across health systems. |
| Ensure follow-through after testing | Structured return-of-results process, provider-facing results letter, referral assistance, and follow-up | Connect relatives with positive results to appropriate preventive care and support. |

The resulting model begins when a clinician identifies an eligible proband and, with consent, refers a proband with a positive genetic result to the Family Heart Foundation. The coordinating organization then obtains a family history, documents outreach preferences, contacts and educates relatives, facilitates genetic and lipid testing, returns results, and helps relatives with positive results connect to preventive care. A detailed operational blueprint and the planned materials are presented in the Supplementary Material.

## Discussion

### Principal findings

In interviews with patients and clinicians, barriers to centralized FH cascade screening extended beyond willingness to undergo testing. Participants described an interconnected sequence of challenges: fragmented access to screening and treatment, uncertainty about privacy and data sharing, distrust of unfamiliar outreach, complicated family relationships, competing demands, clinician time constraints, and limited genetic-counseling capacity. These findings identify the conditions that a centralized program must address before prospective evaluation.

The central implication is that coordination alone is insufficient unless it is paired with trust, flexibility, and continuity of care. Centralized outreach could reduce clinician burden and improve consistency, but it could also be perceived as intrusive or fraudulent if relatives are not prepared for contact. Proband authorization and advance notification, clear program identity, transparent privacy information, and multiple communication pathways are therefore core design requirements rather than optional enhancements.

### Relationship to previous work

Prior U.S. initiatives have used digital family-communication tools, automated reminders, and chatbots to support cascade screening.^12,13^ Our findings help explain why engagement with such tools may vary: technology can simplify outreach, but it does not by itself overcome privacy concerns, low perceived relevance, strained family relationships, language needs, or limited access to follow-up care. Technology-enabled pathways should therefore coexist with telephone, mail, and proband-led options.

Other cascade-screening studies have evaluated health-system identification, hybrid implementation strategies, and pragmatic testing models.^15,16^ The present study adds patient and clinician perspectives on a model that delegates substantial coordination to an external organization. Participants supported centralization chiefly when it reduced work for clinicians and families while preserving consent, choice, and trusted communication.

### Implications for implementation

The findings support several priorities for a future pilot. First, implementation should minimize the number of additional tasks required of clinicians and clearly distinguish responsibilities of the clinic and coordinating organization. Second, probands should control whether and how the program contacts each relative. Third, materials and outreach should be culturally and linguistically responsive and available through multiple channels. Fourth, programs should address the practical consequences of diagnosis by helping relatives understand costs, locate testing, interpret results, and connect to treatment. Finally, privacy protections and data flows should be explicit to patients, relatives, and clinicians.

Prospective evaluation should determine whether these adaptations improve reach without compromising trust or creating new inequities. Relevant outcomes include the proportions of eligible probands and relatives identified, contacted, tested, and linked to care; reasons for nonparticipation; patient and clinician experience; time and cost; and variation in reach across demographic and health-system groups. The current interview study informed the program design but did not test these outcomes.

### Strengths and limitations

A strength of this study was the inclusion of both patients and clinicians, allowing comparison of family, clinical, and health-system perspectives. The advisory panel added expertise in FH, implementation science, patient experience, law, and insurance and enabled direct translation of findings into program components.

The study also has limitations. Participants were recruited primarily from a geographically and institutionally narrow set of sites in Dallas, Texas, and the findings may not represent patients, families, or clinicians in other regions or health systems. Convenience sampling may have favored participants who were more engaged with FH care. The clinician sample was small and intentionally included specialists with experience in FH, which may limit transferability to general clinical settings. Primary qualitative data were collected from patients and clinicians but not from relatives who had not participated in screening, health-system administrators, or payers. The study identified anticipated barriers to a proposed program; it did not evaluate actual program uptake, feasibility, effectiveness, cost, or equity. The resulting blueprint should therefore be considered a testable implementation model rather than an established intervention.

## Conclusions

Patients and clinicians identified multilevel determinants of centralized FH cascade screening, including affordability and access, privacy and trust, family communication, clinician capacity, and continuity of care. These findings informed concrete design requirements for a U.S. centralized model. Prospective testing is needed to determine whether the model improves family reach, testing, and linkage to preventive care.

## Data Availability

All data produced in the present study are available upon reasonable request to the authors

## Supplementary Material

The material below describes the proposed operational model and prospective evaluation. It is presented separately from the completed qualitative study to distinguish interview findings from activities that have not yet been evaluated.

**Supplementary Table S1.**
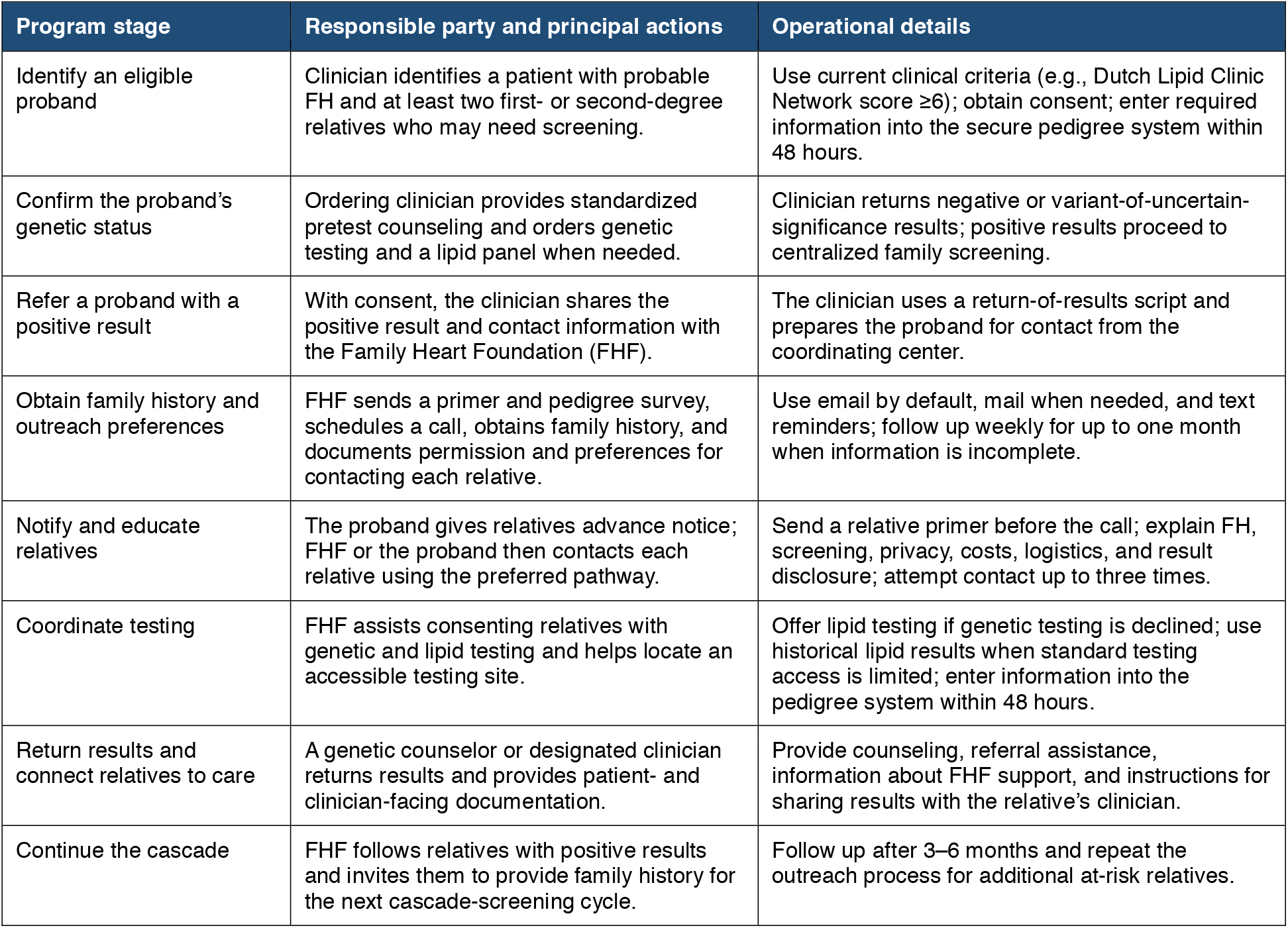
Centralized FH cascade-screening implementation blueprint.

| Program stage | Responsible party and principal actions | Operational details |
| --- | --- | --- |
| Identify an eligible proband | Clinician identifies a patient with probable FH and at least two first- or second-degree relatives who may need screening. | Use current clinical criteria (e.g., Dutch Lipid Clinic Network score $\geq 6$); obtain consent; enter required information into the secure pedigree system within 48 hours. |
| Confirm the proband's genetic status | Ordering clinician provides standardized pretest counseling and orders genetic testing and a lipid panel when needed. | Clinician returns negative or variant-of-uncertain-significance results; positive results proceed to centralized family screening. |
| Refer a proband with a positive result | With consent, the clinician shares the positive result and contact information with the Family Heart Foundation (FHF). | The clinician uses a return-of-results script and prepares the proband for contact from the coordinating center. |
| Obtain family history and outreach preferences | FHF sends a primer and pedigree survey, schedules a call, obtains family history, and documents permission and preferences for contacting each relative. | Use email by default, mail when needed, and text reminders; follow up weekly for up to one month when information is incomplete. |
| Notify and educate relatives | The proband gives relatives advance notice; FHF or the proband then contacts each relative using the preferred pathway. | Send a relative primer before the call; explain FH, screening, privacy, costs, logistics, and result disclosure; attempt contact up to three times. |
| Coordinate testing | FHF assists consenting relatives with genetic and lipid testing and helps locate an accessible testing site. | Offer lipid testing if genetic testing is declined; use historical lipid results when standard testing access is limited; enter information into the pedigree system within 48 hours. |
| Return results and connect relatives to care | A genetic counselor or designated clinician returns results and provides patient- and clinician-facing documentation. | Provide counseling, referral assistance, information about FHF support, and instructions for sharing results with the relative's clinician. |
| Continue the cascade | FHF follows relatives with positive results and invites them to provide family history for the next cascade-screening cycle. | Follow up after 3–6 months and repeat the outreach process for additional at-risk relatives. |

## Supplementary Methods S1. Planned pilot evaluation

The research team developed a prospective evaluation plan to test the feasibility of the implementation blueprint. The planned pilot will recruit 100 unrelated probands with FH from specialty lipid clinics for genetic confirmation. For each proband with a positive genetic result (expected minimum n=50), the Family Heart Foundation will coordinate family screening using the implementation blueprint. The primary outcome will be reach, measured by the numbers and proportions of probands and relatives identified, contacted, tested, and connected to treatment.

Additional planned outcomes include cholesterol measures, FH knowledge, patient experience, implementation barriers and facilitators, program costs, and cost-effectiveness. The evaluation will also include interviews with 20 cascade-screening participants from the two pilot sites and four study staff. These activities are planned and were not part of the completed interview study reported in the main manuscript.

## Supplementary Methods S2. Planned program materials

The interview findings and advisory-panel recommendations informed development of: (1) pretest and post-test scripts for ordering clinicians; (2) a clinician return-of-results script that prepares probands for contact by the Family Heart Foundation; (3) outreach scripts for probands and relatives; (4) a proband primer and sample language for notifying relatives; (5) written primers and letters that can be shared by mail or email; (6) a return-of-results script for relatives; and (7) patient- and clinician-facing result letters. Materials address FH, the purpose and process of cascade screening, testing options, costs, genetic privacy, treatment access, and sources of follow-up support.

