## Appendix 1 for "Patient and Clinician Perspectives on Centralized Cascade Screening for Familial Hypercholesterolemia in the United States: A Qualitative Implementation Study"

### Patient Interview Guide

[Introduction]
**Greeting:** Hello, my name is _________ and I work with [University]. I really appreciate you taking the time to participate in this interview. You are being asked to participate in this study funded by the Department of Defense **because of your unique perspective as someone who has been diagnosed with Familial Hypercholesterolemia or FH.**

**Purpose:** I am part of a research team working with UNC and UT Southwestern on a project to improve screening for Familial Hypercholesterolemia or FH in the US. We are interested in what is important to you as a person with FH. Since FH runs in families, it is important to tell other family members that you have this condition so that they can also get screened for FH. We are planning to interview 10 providers and 20 patients for this study. This study aims to improve the process for screening in families with FH.

**Confidentiality and Introduction:** To start, I'd like to stress that our team will keep everything said here today confidential. Also, nothing you say will be connected with your name. I hope that you will feel free to speak openly.

Please know that there is no right or wrong answer to these questions. Our main goal is to learn from you and have you feel comfortable sharing your experiences, impressions, and beliefs. Our discussion today will last about 30-45 minutes.

Before we begin, I would like to state that the conversation is being audio-taped to help us remember what is said during this discussion. You may ask me to turn off the recorder at any time or simply say you do not want to answer a question. We will compensate you $50 for your time.

**[Section 1] Establishing the Context for the Upcoming Discussion (5 minutes)**

1. Let's start by talking a little about you and your diagnosis. When were you first diagnosed with FH?
    - How old were you at the time?

2. How were you diagnosed?

3. What was your initial reaction when you found out about this diagnosis?
    - How did you feel (angry, happy, etc.)?

4. Do you have children? If yes, how did this affect how you felt about getting your diagnosis?

**[Section 2] Patient History and Experiences (15 minutes)**

5. What do you remember your doctor telling you about FH risk for your family members?

6. Have you shared your diagnosis with any of your blood related siblings and/or parents?
    - Who did you tell?
    - What was this process like for you? How did it go? How was it hard/easy?

- What did you tell them?

7. Have you shared your diagnosis with any of your blood related children (if age appropriate)?
    - Have you had your children screened?
    - What was this process like for you? How did it go? How was it hard/easy?
    - What did you tell them?

8. Have any of your relatives been screened for FH?
    - Have any of your relatives told you that they are being treated for high cholesterol?

9. What factors, if any, might make it difficult to connect your relatives to FH screening?

10. What are things that have helped you connect your relatives to FH screening?

**<u>[Section 3] Patient Attitudes and Beliefs</u> (10 minutes)**
**[Interviewer:** *For this next part of our interview, I would like to share some information about a screening program that we are working on and get your thoughts, ideas, and reactions*].

<u>Brief Description of the Cascade Screening Program:</u> We are planning a cascade screening program. In this program, a patient like you who is newly diagnosed with FH would give permission for their health care provider to notify the Family Heart foundation (which is a patient-driven nonprofit focused on advocacy, research and education of Familial Hypercholesterolemia and elevated Lipoprotein(a), which is sometimes called LP little A) that their patient has been diagnosed with FH and provide them with the patient's contact information. The Family Heart foundation will (1) contact the referred patient to collect details about family, (2) contact relatives and tell them how to get screened.

11. What are your initial impressions upon hearing about this type of screening model?

12. What questions would you have about this type of program?

13. How open would you be to being a part of such a program?

14. What impact do you think the program would have on whether members of your family engage in FH screening?

15. What factors, if any, could make it challenging for you to consent to having the Family Heart Foundation contact your family members? (e.g., concerns about genetic privacy, family dynamics)

16. What might make it challenging for your family members to participate in such a program? (e.g., distance to healthcare)

**<u>[Section 4] Patient Thoughts on Implementing the Centralized Coordinating Model into Daily Practice</u> (20 minutes)**

17. What resources or supports would you want before agreeing to share your family information with the Family Heart Foundation so that they could contact your relatives?
    - educational information?
    - scripts for what to say to relatives?

- information sheet to send to relatives, information about genetic privacy protections, such as the Genetic Information Non-Discrimination Act), which is a law that prohibits health insurance or employment discrimination on the basis of genetic information?
- [If mentioned above] What resources or information would make you feel more comfortable about your genetic privacy?

18. What kind of impact, if any would your participation in a program like this have on the way you talk with relatives about FH?
    - Probe: Would it change how easy or difficult it is to have conversations about FH with family members?

19. What do you think would be the personal benefits of participating in a program like this?

**<u>[Section 5] Closing</u> (5 minutes)**

20. From our discussion today, what are the two takeaways you would emphasize bringing back to the research team as they begin to develop this cascade screening program to help patients and families with FH?

21. Any final thoughts?

*Thank you so much for your time and effort. Following the interview, we will send you a link to your Gift Card*
