## Appendix 2 for "Patient and Clinician Perspectives on Centralized Cascade Screening for Familial Hypercholesterolemia in the United States: A Qualitative Implementation Study"

### Provider Interview Guide

**Pre-Survey Survey Questions:**

1. **Gender**
2. **Age**
3. **Years since completing training**
4. **# FH patients per month**

**Implementing Cascade Screening for Familial Hypercholesterolemia in the US**
**Provider Semi-Structured Interview Questions**
**Megan Roberts (PI)**

[Introduction]
**Greeting:** Hello, my name is _________ and I work with the University of North Carolina at Chapel Hill. I really appreciate you taking the time to participate in this interview. You are being asked to participate in this study **because you have direct involvement with patient care.**

**Purpose:** I am part of a team working with **UNC and UT Southwestern on a project to learn more about how to improve screening for Familial Hypercholesterolemia or FH in the US**. In particular we are interested in improving processes for cascade screening and creating a systematic process of contacting and screening relatives of someone who has been diagnosed with FH.

We are interested in what is important to healthcare providers like you and potential ways to improve how something like this could be integrated into clinical practice. We are planning to interview 10 providers and 20 patients for this research project. The information that you provide will assist us in optimizing our process for centralized cascade screening for FH.

**Confidentiality and Introduction:** To start, I'd like to stress that our team will keep everything said here today confidential. Also, nothing you say will be connected with your name. I hope that you will feel free to speak openly.

Please know that there is no right or wrong answer to these questions. Our main goal is to learn from you and have you feel comfortable sharing your experiences, impressions, and beliefs. Our discussion today will last about 30-45 minutes and following the interview we will compensate you $50 for your time.

Before we begin, I would like to state that the conversation is being audio-taped to help us remember what is said during this discussion. You may ask me to turn off the recorder at any time or simply say you do not want to answer a question.

**[Section 1] Establishing the Context for the Upcoming Discussion (5 minutes)**

1. Tell me about your practice. What type of patient population do you see?

2. How much of your practice involves diagnosing and treating patients with FH?

**[Section 2] Provider Experience with Cascade Screening for FH (10 minutes)**

3. I'm interested to know what your process is when a patient is newly diagnosed with FH.

    a. Do you refer your patients to genetic counseling? If so, when (e.g., prior to genetic testing, following a diagnosis)?

b. Walk me through your process for offering cascade screening to the patient's genetic relatives.

4. From your perspective, what barriers currently exist for obtaining family history information to support cascade screening efforts?

5. From your perspective, what facilitators are in place that help you currently obtain family history information to support cascade screening efforts?

**[Section 3] Provider History and Knowledge with the Centralized Coordinating Model (10 minutes)**
*[Interviewer: Now I would like to ask you some questions about our proposed centralized screening program, which is modeled after the Dutch government-funded cascade screening program. In The Netherlands, this program has helped identify individuals with FH early in life and led to improved treatment and reduced cardiovascular disease. A key aspect of the Dutch model was a centralized office that contacted patients and coordinated efforts. We aim to implement a similar model here in the US, in partnership with the Family Heart Foundation, which will serve as the coordinating center. Under this model, providers would inform their patients who are newly diagnosed with FH that the Family Heart Foundation would be contacting them. The provider's clinic would notify the Family Heart foundation that a patient has been diagnosed with FH and provide them with the patient's contact information. The Family Heart Foundation will (1) contact the patient to collect details about family, and (2) contact relatives and tell them how to get screened.*

6. What are your initial impressions upon hearing about this type of screening model?

7. What questions would you, as a provider, have about this type of program?

8. In general, **how receptive would you be to** introducing the Centralized Coordinating screening model in your clinic to your patients? **[Beliefs about the Intervention]**

9. What impact, if any, could the Centralized Coordinating screening model have on patients?
    a. Probe: Physically? Mentally? Socially? **[Beliefs about the Intervention]**

10. From your perspective, what is the best way for the Family Heart foundation to receive information about a newly diagnosed patient with FH who needs cascade screening? [e.g., local provider enters patient into a HIPAA compliant website] **[Cross-cutting]**

**[Section 4] Provider Thoughts on Implementing the Centralized Coordinating Model into Daily Practice (15 minutes)**

11. What barriers would you anticipate to implementing the Centralized Coordinating screening program in your clinical setting? (Patient level, provider level, practice level) **[Cross-cutting]**

12. Who would be the key influential individuals to get on board with implementation of the Centralized Coordinating screening program in your clinic? **[Processes: Engaging]**

    a. Anyone else who would be involved in the implementation? If so, who?

13. How well does the Centralized Coordinating screening program seem to fit with existing work processes and practices in your clinical setting? **[Inner Setting]**

    a. What kinds of infrastructure changes would be needed to accommodate the Centralized Coordinating screening program in your clinic? **[Inner Setting]**

b. What changes do you think you would need to make to the Centralized Coordinating screening program itself so it would work effectively in your clinical setting? **[Compatibility]**

14. What resources (e.g., time, money, training, education, physical space) and supports (e.g., clinical decision support, educational meeting, information about genetic privacy, toolkits) would you need to successfully implement and administer the Centralized Coordinating screening program in your clinic? **[Available Resources; Design Quality and Packaging]**

15. What kind of metrics do you think implementers should collect to evaluate the effectiveness of implementing the Centralized Coordinating screening program? **[Evaluating]**

**[Section 5] Closing (5 minutes)**

16. From our discussion today, what are two takeaways you would emphasize bringing back to the research team as they begin to develop an intervention to improve cascade screening for FH?

17. Any final thoughts?

*Thank you so much for your time and effort. Following the interview, we will send you a link to your Gift Card*
