## Appendix 3 for "Patient and Clinician Perspectives on Centralized Cascade Screening for Familial Hypercholesterolemia in the United States: A Qualitative Implementation Study"

### Analysis Codebook

| Code | Description | Data Source(s) |
|---|---|---|
| **Section 1. Participant Background** | | |
| Patient population and FH | Include comments about the practice, types of patient population seen, and how much of the practice involves diagnosing and treating patients with FH. | Providers |
| **Section 2. Provider Experience** | | |
| Process for FH diagnosis<br><br>(WORKFLOW) | Include comments about the diagnosis process for a newly diagnosed patient with FH, any comments about referring patients to genetic counseling and if so, when (e.g., prior to genetic testing, following a diagnosis). | Providers |
| Process for Cascade Screening (WORKFLOW) | Include comments about the process for offering cascade screening to the patient's genetic relatives. | Providers |
| Family History Info (WORKFLOW) (ED MATERIALS) | Include comments about barriers and facilitators that exist for obtaining family history information to support cascade screening efforts. | Providers |
| **Section 3. Patient Experience** | | |
| FH Communication with doctors (WORKFLOW) (ED MATERIALS) | Include comments about what the doctor communicated about FH risk for family members. Also include comments if participants say they cannot remember the communication. | Patients |
| FH Communication with relatives (WORKFLOW) (ED MATERIALS) | Include comments about whether patients shared their diagnosis with any blood related children, siblings and/or parents, who they told, what was this process like, how it went, and what was communicated. Include comments related to discussions the patients have had with their relatives about elevated cholesterol and FH. Also include comments/reasons shared for why participant did not communicate with relatives. | Patients |
| FH Screening by relatives (WORKFLOW) (ED MATERIALS) | Include comments about whether any relatives have been screened for FH, any comments about relatives sharing that they have been screened for, diagnosed with, and/or are being treated for high cholesterol. | Patients |
| FH Screening Barriers (WORKFLOW) (ED MATERIALS) | Include comments about factors that might make it difficult to connect relatives to FH screening. | Patients |
| FH Screening Facilitators (WORKFLOW) (ED MATERIALS) | Include comments about factors that have helped connect relatives to FH screening. | Patients |
| **Section 4. Provider and Patients: Centralized Coordinating Model (CCM)** | | |
| Family Heart Foundation (MODEL) | Include comments about the best way for the Family Heart Foundation to receive information about a newly diagnosed patient with FH who needs cascade screening. [e.g., local provider enters patient into a HIPAA compliant website]. [Cross-cutting] | Providers |

| Code | Description | Data Source(s) |
|---|---|---|
| Impact on family participation (MODEL) | Include comments about any impact (positive or negative) CCM would have on whether family members may engage in FH screening. | Patients |
| Impact on family discussions (MODEL) (ED MATERIALS) | Include comments about any impact a patient's participation in a program like this may have on the way they talk with relatives about FH. Include comments if it would change how easy or difficult it is to have conversations about FH with family members. | Patients |
| Providing consent to contact (MODEL) | Include comments about participant's comfort level for consenting to having the Family Heart Foundation contact their family members. Also include comments about any issues that should be considered as related to getting consent (e.g., concerns about genetic privacy, family dynamics,) or considerations (e.g. patient should notify family first before cold contact) | Patients |
| Barriers to participation (MODEL) | Include comments about factors that could make it challenging for your family members to participate in such a program (e.g., distance to healthcare, skeptical of initial phone contact, lack of interest). | Patients |
| Openness and receptiveness to CCM (MODEL) | Include comments about **degree of openness** to being a part of such a program. (For providers) Also include **receptiveness** to introducing the Centralized Coordinating screening model in your clinic to your patients. [Beliefs about the Intervention] | Providers Patients |
| CCM questions (MODEL) | Include comments about specific questions about this type of program brought up by participants. Also include comments indicating that participants have no questions. | Providers Patients |
| CCM impact on patients (MODEL) | Include comments about any impact (i.e. physically, mentally, socially) CCM may have on patients. Include comments about the personal benefits of participating in a program like this for patients and/or their family members. Also include any comments about the way that CCM could negatively impact patients. [Beliefs about the Intervention] | Providers Patients |
| **Section 5. Provider and Patients: Implementing the Centralized Coordinating Model** | | |
| Implementation barriers and concerns (MODEL) | Include comments about any anticipated barriers to implementing CCM in the clinical setting (Patient level, provider level, practice level.) [Cross-cutting] | Provider |
| Implementation Partners (MODEL) | Include comments about the key influential individuals to get on board with implementation of CCM in your clinic. Also include discussions about the staff needed to implement CCM. [Processes: Engaging] | Provider |
| Existing work processes (MODEL) (WORKFLOW) | Include comments about how well the CCM seems to **fit** with existing work processes and practices in the clinical setting? [Inner Setting]. Also include comments about the kinds of infrastructure changes that would be needed to accommodate CCM in the clinic? [Inner Setting], and any changes needed to make to CCM itself so it would work effectively in the clinical setting? [Compatibility] | Provider |
| Implementation metrics (WORKFLOW) | Include comments about the kind of metrics implementers should collect to evaluate the effectiveness of implementing the Centralized Coordinating screening program. [Evaluating] | Providers |
| Implementation resources needed (WORKFLOW) | Include comments about resources (e.g., time, money, training, education, physical space) and supports (e.g., clinical decision support, educational meeting, information about genetic privacy, toolkits) need to successfully implement and administer the CCM in the clinic. [Available Resources; Design Quality and Packaging] | Providers Patients |

| Code | Description | Data Source(s) |
| --- | --- | --- |
| | Note: Comments about staff needed to implement CCM should be coded as Implementation Partners—NOT Implementation resources needed.<br><br>Also include comments about resources or supports patients would want before agreeing to share family information with the Family Heart Foundation so that they could contact relatives or things patients would want to make them feel more comfortable about their genetic privacy. | |
| Suggestions and Recommendations | Include suggestions, recommendations, or key points to emphasize and bring back to the research team as they begin to develop an intervention to improve cascade screening for FH. | Providers<br>Patients |
