## Appendix 5 for "Patient and Clinician Perspectives on Centralized Cascade Screening for Familial Hypercholesterolemia in the United States: A Qualitative Implementation Study"

Post-test Counseling (Known FH Diagnosis with pathogenic/likely pathogenic variant identified DISCOVER FH)

**Results: POSITIVE**

A ***likely pathogenic (disease causing) variant was identified in ***gene. This variant is known to cause familial hypercholesterolemia and explains your history of familial hypercholesterolemia.

**Inheritance:**

Heterozygous familial hypercholesterolemia (HeFH) is inherited in an autosomal dominant fashion from parent to child. The ***likely pathogenic variant found in you was likely passed down to you from your mother OR your father. Everyone has two copies of the *** gene, one you inherit from your mother and one from your father. Having a variant in one copy of your ***gene will cause familial hypercholesterolemia.

**HoFH?

**Variant details:**

Gene:

Variant:

Lab:

Test requisition number:

**Implications for proband: You will require life-long lipid lowering therapy which can substantially reduce your risk of a future cardiovascular event.**

**Implications for family members:**

Heterozygous FH (HeFH) is inherited in an autosomal dominant manner. Almost all affected individuals inherit the familial mutation from an affected parent. There is a 50% chance that you inherited this from your mother and a 50% chance you inherited this from your father. Additionally, children and full siblings (those that share the same mother and father) have a 50% chance of inheriting this same variant and should have testing. More distant relatives (half siblings, aunts, uncles, grandparents, cousins) are also at risk and should consider screening for familial hypercholesterolemia.

**Next steps:**

Your contact information will be shared with the Family Heart Foundation, ***FHF brief description?***. Someone from the Family Heart Foundation will contact you ***by modality*** within X days to discuss screening for your family members.
