## Appendix 7 for "Patient and Clinician Perspectives on Centralized Cascade Screening for Familial Hypercholesterolemia in the United States: A Qualitative Implementation Study"

**Bullet Points for Proband Call**

Thank you for taking my call and for participating in DISCOVER FH

First, let me ask if you have any questions about your diagnosis of FH or your own treatment goals.

The goal of DISCOVER FH is to encourage **cascade screening** in your family.

Cascade screening involves testing your family members, both children and adults to find others who may have FH and allows them to get treatment early in life to reduce their risk of heart disease and stroke.

Unfortunately, in the United States we have not been very successful in implementing cascade screening. We think this is because we have left it up to individuals who are diagnosed with FH to inform their relatives that they are at risk.

The Netherlands, however, had one of the most successful cascade screening programs. It’s believed 70% of people with FH in the Netherlands were diagnosed through cascade screening. Our program is trying to adapt the Dutch program to the United States.

The Dutch used a centralized coordinating team. When a person, like you for example, was found to have FH, the centralized coordinating team reached out to the person and collected information on his/her family. Then, a member of the team reached out to each family member and arranged testing.

This is where we at the Family Heart Foundation fits in, we can reach out to each of your family members on your behalf to explain FH and arrange for free and convenient genetic and cholesterol testing. After the testing is complete we will follow-up with your relatives and explain the results and help them find care in their community if they need to find a health care provider.

What we ask of you is that you give each of your first-degree relatives (parents, siblings, children) a heads-up call/text/email about your diagnosis and that someone from the Family Heart Foundation will be calling/emailing/texting them to explain how your diagnosis of FH potentially impacts them. We have provided you with a potential script that you can use when talking to your relatives. It is in your information packet.

We ask that you give us the names and best contact information for all your first-degree relatives (parents, siblings, children).

We will send each of them an information packet that will include information about FH, the DISCOVER FH program and a letter that they can share with their primary care provider.

We will wait a few days after sending the information packet and then we will contact your relatives to arrange testing.

Do you have a list of names ready for us today?
