## Appendix 8 for "Patient and Clinician Perspectives on Centralized Cascade Screening for Familial Hypercholesterolemia in the United States: A Qualitative Implementation Study"

**Bullet Points/Script for Relative Call**

**Answering Machine**

This is a message for <<RelativeName>>. This is <<Care Navigator>>,calling from The Family Heart Foundation with information that could be important to your health. You should have also received information from us via<<mail/email >>. I will try to call back at another time, or you may call me at<<Insert Direct Line>>.If you leave a message for us, please include the best time to contact you. Thanks, and have a great day.

**Call Answered:**

<Confirm identity>

Introduction:

I am a Care Navigator with the Family Heart Foundation. We’re working with (Proband’s) healthcare providers to help share some important health information they learned recently. You should have received some information about this via <Mail/email>.

<Check if received packet>

I am calling you because your (brother, sister, mother, father, daughter, son) (relative’s name) was diagnosed with familial hypercholesterolemia (FH for short) through a program at the University of Texas South Western (UTSW) called DISCOVER FH. Our team at the Family Heart Foundation is partnering with UTSW/Dallas VA to reach out to (relatives’ name) family members to explain what their diagnosis means for you, what next steps you can take to protect your health and the health of your family members.

Has (relatives name) been in touch with you about this program?

Introduce self: My name is <<Care Navigator>> and I work with the Family Heart Foundation. I’m a <<genetic counselor>> with the Family Heart Foundation, which is a non-profit patient research and advocacy organization. I have special training in working with families like yours to help provide personalized help and support need as they make decisions about their genetic health.

Let me stop here and ask if you have any questions about FH or about this family screening program?

**Information about FH:**

FH is a serious genetic condition that affects about 1 in 250 people. And it runs in families, so because (relative’s name) has FH, each of his/her first-degree relatives (parents, siblings, and children) has a 50% chance of having FH.

People with FH have very high low-density lipoprotein-cholesterol (LDL-C), also known as “bad cholesterol.” The LDL-C in people with FH is high from birth, so they are at high risk for heart attack and stroke at a young age.

LDL-C deposits in the arteries that carry blood to the brain and heart. This leads to blockages that can cause heart attacks, the need for stents, bypass surgery, stroke, and even death. **Fortunately, treatment with cholesterol-lowering medications can significantly reduce this risk.**

FH is very common for an inherited condition, but most do not know they have it. They might think they just have high cholesterol. Often, even doctors miss the FH diagnosis. Recent studies have shown that by age 40, a quarter of untreated adults with FH will have had a cardiac event like a heart attack or stroke and 7% will have passed away of cardiac disease.

Because people with FH have such a high risk of heart attack and because their cholesterol level is much higher than people who have high cholesterol for other reasons, we often treat their high cholesterol differently than high cholesterol caused only by other things like diet or lifestyle factors.

It’s important that you be tested for FH for your own health. And if you learn that you have FH, you can also alert your other family members to their own FH risks. People are born with FH, and the high cholesterol levels start at birth, so it is very important for younger generations in your family to find out as young as possible if they have FH. Cascade testing for FH is recommended as young as age 2 years, and treatment for FH is recommended to start as young as 8-10 years of age. This will give the children in your family the best chance to not develop heart disease and stroke. You can help protect your family.

**Information about family screening:**

Family screening, also known as cascade screening, involves testing family members of a person diagnosed with FH, both children and adults, to find others who may have FH. This allows them to get treatment early in life to reduce their risk of heart disease and stroke.

The purpose of my call today is to offer to arrange free testing including genetic testing and cholesterol testing. The genetic testing will be through GENinCode a genetic testing laboratory and the cholesterol testing which will include both a lipid panel and a lipoprotein (a) will be through Quest laboratories.

**Genetic testing:**

Because we know the specific genetic cause of <<*Proband Name*>>’s FH, a genetic test is a clear way to determine if you have inherited FH. It can provide you and your family members with an accurate diagnosis based on <<*Proband Name*>>’s result. Learning if you have FH can help you get the best treatment possible.

Genetic testing uses a simple saliva test that can give you a yes/no answer to whether you have the same genetic variant or mutation <<proband>> has. The saliva test is through a genetic testing company called GENinCode. The test is offered at no-cost and would be sent directly to your home for you to complete and return.

Additionally, this genetic test includes what is known as a polygenic risk score for high LDL. While your relative has a single variant or mutation that is causing their FH, there are other genetic changes which also contribute to a person’s overall risk for having high LDL cholesterol. It is possible that you may not have FH, but still have genetically elevated LDL cholesterol. This information may be useful for guiding your health care and will be discussed at the time of your results.

Discuss potential for alternative monogenic explanations for FH.

**Cholesterol testing:**

In addition to the genetic testing, we are also offering free cholesterol testing (lipid testing) through Quest. A cholesterol test is a simple blood test that can also help your healthcare provider find out if you have FH. Having routine cholesterol tests can help your healthcare provider monitor your cholesterol levels and estimate your risk for things like heart attack and stroke. If your cholesterol levels are high, your healthcare provider may prescribe a cholesterol-lowering medication and discuss heart healthy lifestyle choices with you to help reduce your risk of heart attack and stroke.

Even if you’ve had recent cholesterol testing, we would appreciate an up-to-date test to make sure that everybody in this study has labs completed from the same laboratory.

<<Mail Logistics>>

<<Collect relevant information>>

- Personal history of ASCVD
- Medication history?
- Additional family history as needed

<<Follow up plan>>

- Plan to complete genetic test by XYZ
- Schedule lipid testing on call

Alternatives: If not interested in participating in program

- Discuss reasons for disinterest
  - Address any concerns/misconceptions as needed
  - Interested in potential f/u interview to help us better understand reasons they are not interested?
- Discuss previous lipid testing if available
- Discuss sharing this information with their health care provider. Offer to share letter they can take into their HCP

Results review:

Once we have your cholesterol test results and your genetic testing results either myself or an FH expert from our team will set up a call to review these results with you. We will discuss your FH status, your risk, and next steps. We can help you find a lipid specialist near you to find the right treatment if needed. We can also help you share this information with your primary care provider.

If you are positive, we will discuss any additional cascade testing of your relatives if indicated (Children, other siblings, etc.)

General Discussion/Talking Points:

- High cholesterol vs FH
  - High cholesterol is common, may or may not be related to FH
  - Treated differently, importance of early and aggressive treatment for FH
- Treatment if positive
  - Discuss standard of care for treatment
  - FH not responsive to dietary and lifestyle interventions
- Address cost concerns
  - All testing will be covered as part of the study
  - Can assist in finding right care after results are in
  - Opportunity to review results with an FH expert at no cost after to receiving results
- Frame as health benefit for family members
  - Testing can be informative for your children’s health
- Address perceived barriers such as insurance concerns (e.g., below)
  - Review GINA
  - Life/Long term care/ disability insurance prior to testing if interested
