## Appendix 9 for "Patient and Clinician Perspectives on Centralized Cascade Screening for Familial Hypercholesterolemia in the United States: A Qualitative Implementation Study"

### DIrect SCreening Of RelatiVEs to Reveal FH

Thank you for agreeing to be part of the DISCOVER FH Study. This letter is meant to provide information for you regarding your recent testing results as well as details about our cascade Family Screening program to help your relatives get the care they need.

##### Summary of Your Results

Based on your recent genetic testing results, you were found to have familial hypercholesterolemia (FH). A copy of your genetic test report is included in this packet.

##### What is Familial Hypercholesterolemia?

Familial hypercholesterolemia (FH) is a common genetic condition that causes very high low-density lipoprotein (LDL) cholesterol or "bad cholesterol." If left untreated FH leads to early heart attacks and strokes. FH is inherited, meaning it is passed down from parents to children.

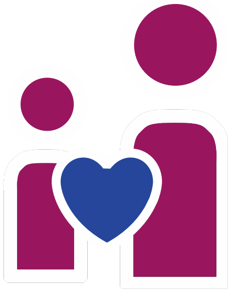

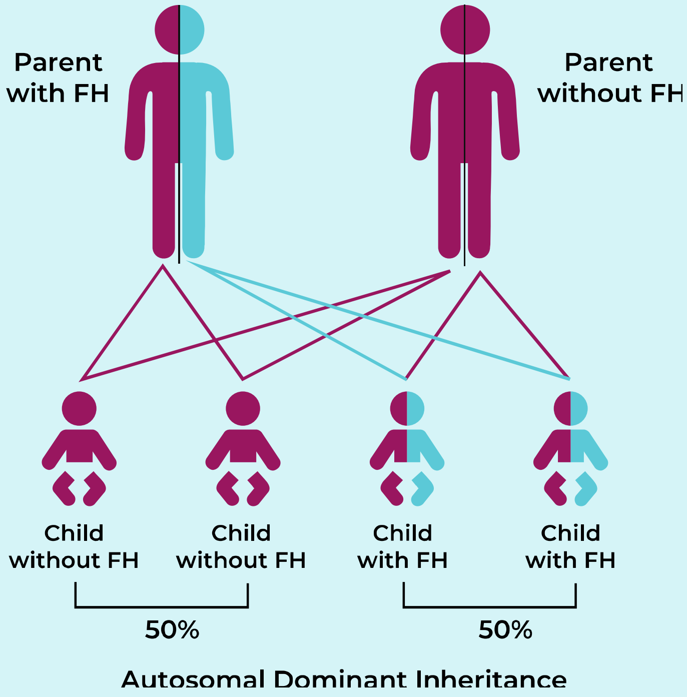


##### How is Familial Hypercholesterolemia Inherited?

Everybody has 2 copies of each gene. You get one copy of a gene from your mother and one copy of a gene from your father. Having a single disease-causing variant (also known as genetic changes, or mutation) in one copy of a gene is enough to cause FH. This is what's known as dominant inheritance. Your parents, siblings, and children each have a 50% chance of having the same FH causing genetic variant as you. More distant relatives may also be at risk and should consider screening.

#### What is Cascade Screening?

Cascade screening or family screening, is the process of testing at risk relatives of someone known to have the condition. Once we know an individual has FH, like yourself, all at risk relatives should be screened using genetic testing and cholesterol testing. This will give your close relatives a yes/no answer as to whether they also have FH. If they are found to also have FH, the process then extends (or cascades) to their close relatives. For example, if you have FH and a sibling is also found to have FH, then your sibling's children would be offered testing.

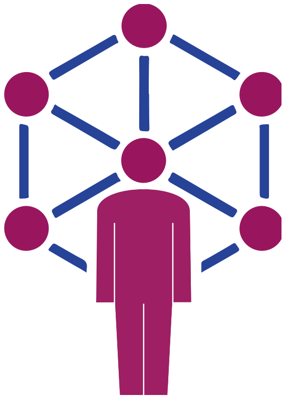

#### How Does Cascade Screening Work in DISCOVER FH?

An expert from the Family Heart Foundation will reach out to you to discuss cascade screening. This call will include a review of familial hypercholesterolemia, an overview of the cascade screening process and a review of your family history. With your permission, the expert will collect contact information for your relatives and develop a plan for contacting your at-risk relatives.

You will be asked to give your relative(s) a "heads up" message to let them know the Family Heart Foundation will be contacting them. See below for examples of messages to use with your family members.

Someone from the Family Heart Foundation will also send basic information to your relatives about FH and this DISCOVER FH screening program.

The expert you are working with at Family Heart Foundation will then contact your relatives to discuss FH and their risk in more detail. Free testing will be provided to your relatives. This testing will include at-home saliva test for genetic testing as well as a simple blood test to check their cholesterol levels. If your relatives need help finding a local lab for this blood test, the Family Heart Foundation will help them find a convenient location. Once results are available, the Family Heart Foundation will review these results at no cost with your relatives and help them share this information with their healthcare providers or help them find a specialist if needed.

#### Which of my relatives would be eligible for family screening?

All your first-degree blood relatives are eligible for this no-cost family screening program through DISCOVER FH. This includes parents, siblings, and children (over age 2). In some circumstances more distant relatives may be eligible for screening through this program.

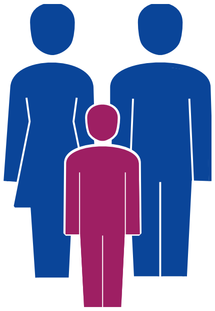

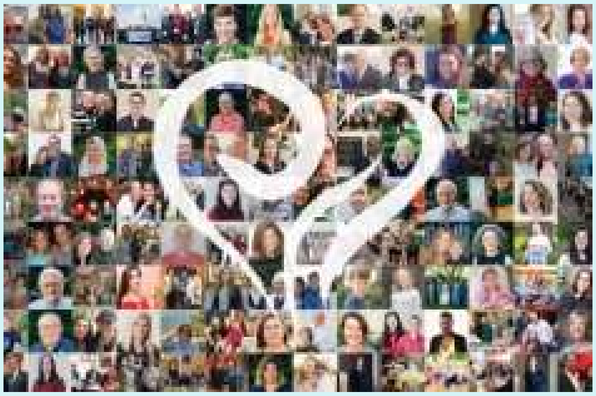

##### Who is the Family Heart Foundation?

The Family Heart Foundation is a non-profit patient research and advocacy organization. They are partnered with UTSW and the Dallas VA to help improve family screening for FH. Learn more at *familyheart.org*

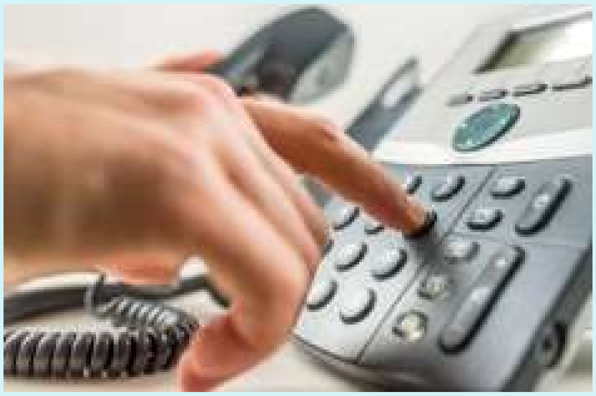

##### What if I have relatives who I don't want you to contact?

We recognize families can be complicated. When you speak with the FH expert at Family Heart Foundation, we can discuss alternatives to participating in this screening program.

#### For questions regarding

##### Your Healthcare

VA | U.S. Department of Veterans Affairs

UTSouthwestern Medical Center

##### The Study

UTSouthwestern Medical Center

***-***-****

<email address>

##### Family Screening

FAMILY HEART FOUNDATION
Lipoprotein(a) & Familial Hypercholesterolemia

Family Heart Foundation

582-203-0755

##### Where can I find additional information about cascade screening?

www.familyheart.org/what-is-family-screening

www.cdc.gov/genomics/disease/cascade_testing/cascade_finding.htm

##### What are my next steps?

You will receive a text message and a phone call from 582-203-0755 to discuss this program in more detail. In the meantime, try to collect the names and contact information (phone numbers, email address, mailing addresses) for all your close relatives.

Thank You

**For participating in DISCOVER FH!**

### Discover FH Family Screening Program

#### How It Works

1. You were found to have a genetic cause for FH
2. An expert from the Family Heart Foundation calls to discuss family screening for your relatives
3. You give your relatives a "heads up" letting them know they will be contacted
4. Your relatives are sent information via email or mail (USPS) by the Family Heart Foundation
5. An expert from the Family Heart Foundation will call your relatives to discuss and coordinate free screening
6. Family Heart Foundation will review results with your relatives and if needed, help them find the care they need

#### Here's What To Say When You Give Your Relative A "Heads Up"

"I learned that I have a serious genetic condition called FH, or inherited high cholesterol. FH runs in families. I want to protect your health and the health of our family members, so I asked an expert at the Family Heart Foundation to contact you."

"The Family Heart Foundation team are experts in FH and can tell you more about it. They can explain the health risks for people like me who have FH, the treatments for FH, and how to get tested at no cost to learn if you have FH."

"The FH expert can also tell you about testing options for FH and give you important information to share with your doctor. They will reach out soon by mail or email (***), first."

### Script for discussing your FH diagnosis with your relatives

Hi (relative's name), I am calling to let you know that I have been diagnosed with familial hypercholesterolemia (also known as FH).

FH is a genetic condition that affects about 1 in 250 people. And it runs in families, so because I have FH, each of my first-degree relatives (parents, siblings, and children) has a 50% chance of having FH.

People with FH have very high low-density lipoprotein-cholesterol (LDL-C), also known as "bad cholesterol." The LDL-C in people with FH is high from birth, so they are at high risk for heart attack and stroke at a young age.

LDL-C deposits in the arteries that carry blood to the brain and heart. This leads to blockages that can cause heart attacks, the need for stents, bypass surgery, stroke, and even death. **Fortunately, treatment with cholesterol-lowering medications can significantly reduce this risk.**

I was diagnosed through a program at the University of Texas Southwestern (UTSW) and the Dallas VA called DISCOVER FH. The team at UTSW and the Dallas VA are partnering with a nonprofit research and advocacy organization called the Family Heart Foundation to help them reach out and offer free genetic testing and cholesterol testing to family members all over the country.

I am calling to let you know that an expert from the Family Heart Foundation will be reaching out to you to talk about FH and work with you to schedule free testing.

Once you have the testing done, the Family Heart Foundation expert will call you to review the results. The Family Heart Foundation will also give you information that you can give your own primary care provider or help you find an FH specialist in your area.

FAMILY HEART FOUNDATION®
Lipoprotein(a) & Familial Hypercholesterolemia

www.familyheart.org
