## Appendix 10 for "Patient and Clinician Perspectives on Centralized Cascade Screening for Familial Hypercholesterolemia in the United States: A Qualitative Implementation Study"

{Today’s Date}

Dear {Relative’s Name},

I hope this letter finds you well. My name is {XXX} and I am a Care Navigator at the Family Heart Foundation working with {Proband’s Name} and their healthcare providers at <<UTSW/Dallas VA>>. {Proband’s Name} asked me to help share some important health information with you that may also affect your health and the health of your other family members.

**{Proband’s Name} was recently diagnosed with a serious genetic disorder called** **Familial Hypercholesterolemia (FH). FH is also known as inherited high cholesterol**. Having FH causes very high LDL (“bad”) cholesterol levels from birth. Having high levels of bad cholesterol puts one at a **higher risk for early heart disease, heart attack, stroke, and possibly even d****eath, *if left untreated*.** Fortunately, there are many effective treatment options available.

As {Proband’s Name}’s {Relation to Proband}, **it is important to understand your own FH risks and options for next steps.**

**FH runs in families.** Based on your relation to {Proband’s Name}, you have a **__% chance** of also having inherited FH. Overall, parents, children, brothers and sisters of people with FH have a 50% chance of also having inherited FH or not. Other family members (aunts, uncles, nieces, nephews, cousins, grandchildren, etc.) may also have inherited this serious genetic disorder.

Since sharing complex health information can be hard, your relative gave me your contact information and asked that I reach out to you directly to discuss this information.

**I will call you soon to follow up on this information.**

**During our call, we will discuss more about:**

- {Proband’s Name}’s FH result.
- What this information means for you.
- How you can get **free testing** to learn if you also have FH
- Next steps you can take to protect your heart health and the health of your family members.

For more information about FH, you can visit **The Family Heart Foundation’s webpage** at [https://familyheart.org/discoverfh](http://www.familyheart.org/discoverfh).

If you do ***not*** wish to hear from me, please contact the study team (toll-free) at ******* or ******* to let me know.

Please feel free to also contact me before I call with any questions or concerns you may have. Otherwise, I will talk to you soon!

Sincerely,

{Signature}
