## Appendix 13 for "Patient and Clinician Perspectives on Centralized Cascade Screening for Familial Hypercholesterolemia in the United States: A Qualitative Implementation Study"

POSITIVE Results Letter to Relative-

Dear _____,

Thank you for taking part in this Research Program involving Cascade Family Screening to Find

Familial Hypercholesterolemia (FH).

It was a pleasure working with you to help arrange your screening for FH. This letter is for your records, and it summarizes the results of your screening. A copy of your test results is included, and you are encouraged to share this with your health care provider.

**Summary:**

Based on your ***screening results, personal history, and family history, it is **probable** that you have familial hypercholesterolemia. It is important that you speak with your health care provider about these results. Early treatment with cholesterol lowering medications are available and they will help lower your risk for heart disease and stroke.

It is known that individuals with FH, if left untreated are at a greater risk for heart attack and stroke.

Lab Results:***

Medications: ***

Personal History: ***

Family History: ***

**About Familial Hypercholesterolemia:**

Familial hypercholesterolemia (FH) is a common genetic condition that causes very high low density lipoprotein (LDL) cholesterol or “bad cholesterol”. FH is inherited, meaning it is passed down from parents to children.

**Next Steps:**

It is important to speak with your health care provider about these results.

There are a number of FH specialists who you might consider seeing as well.

An FH specialist will be able to work with you to determine the right treatment plan for you. The Research Team is here to answer your questions and can help you find an FH specialist if needed.

**General Treatments Goals for FH:**

For people with FH but no history of heart disease or stroke, the goal LDL-C is less than 100 mg/dL.

For people with FH who have already developed heart disease or stroke, the goal is an LDL-C of less than 70 mg/dL. Recent recommendations for LDL-C are pushing for less than 55 mg/dL in higher risk people. In general, the lower the LDL level the better.

**For family members:**

Because you likely have FH, your family members are also at risk of having FH. Close family members (parents, siblings, and children) all have a 50% chance of having FH and should consider screening. If your relatives need help finding screening, we are happy to help them with this.

**Staying Healthy**

In addition to maintaining an acceptable level of blood cholesterol through the use of cholesterol lowering medication

you can help lower your risk of heart disease and other conditions by following a healthy diet, engaging in exercise regularly, maintaining healthy levels of blood sugar and blood pressure, not smoking, and keeping your weight in a healthy range. Your health care provider can provide you with more guidance on these lifestyle measures.

**<<Resources** FamilyHeart.org>>

Sincerely

***

Contact Info

Negative Results Letter to Relative-

Dear _____,

Thank you for taking part in this Research Program involving Cascade Family Screening to Find

Familial Hypercholesterolemia (FH).

It was a pleasure working with you to help arrange your screening for FH. This letter is for your records, and it summarizes the results of your screening. A copy of your test results is included, and you are encouraged to share this with your health care provider.

Based on your ***screening results, personal history, family history, and the medications you regularly take, it is **unlikely** that you have FH.

Lab Results:***

Medications: ***

Personal History: ***

Family History: ***

**What does this mean for family members.**

Because you have a first degree relative who has been diagnosed with FH other first-degree relatives may still be at risk for FH and they should be screened.

Relatives not at risk should still follow population screening guidelines.

Current recommendations are:

Children should be screened with a cholesterol panel between the ages of 9-11 and again between 17-21.

Adults should be screened every 4-6 years.

**Staying Healthy**

You can lower your risk of heart disease by following a healthy diet, engaging in exercise regularly, maintaining healthy levels of blood cholesterol, blood sugar and blood pressure, not smoking, and keeping your weight in a healthy range. Your health care provider can provide you with more guidance on these lifestyle measures.

**<<Resources** FamilyHeart.org>>

Sincerely

***

Contact Info

Genotype(+), Phenotype(-) Results Letter to Relative-

Dear _____,

Thank you for taking part in this Research Program involving Cascade Family Screening to Find

Familial Hypercholesterolemia (FH).

It was a pleasure working with you to help arrange your screening for FH. This letter is for your records, and it summarizes the results of your screening. A copy of your test results are included, and you are encouraged to share this with your health care provider.

**Summary:**

Based on your genetic testing results

(screening results, personal history, and family history)- delete?

you have familial hypercholesterolemia. It is important you speak with your health care provider about these results. Your LDL-C is lower than we typically see with FH, but it still requires treatment. Treatment with cholesterol lowering medications will help lower your risk for heart disease and stroke.

It is known that individuals with FH, if left untreated are at a greater risk for heart attack and stroke.

Lab Results:***- (LDL-C less than 190 mg/dL)

Genetic testing- positive (state identified gene defect: LDLR, ApoB, PCSK9)

Medications: ***

Personal History: ***(no history of Cardiovascular disease)

Family History: *** (no history of early Cardiovascular disease in first degree relative)

**About Familial Hypercholesterolemia:**

Familial hypercholesterolemia (FH) is a common genetic condition that typically causes very high low density lipoprotein (LDL-C) cholesterol or “bad cholesterol”. FH is inherited, meaning it is passed down from parents to children. Even though your LDL-C level is lower than what is seen in FH, your family members may still have a very elevated level.

**Next Steps:**

It is important to speak with your health care provider about these results.

There are a number of FH specialists who you might consider seeing as well.

An FH specialist will be able to work with you to determine the right treatment plan for you. The Research Team is here to answer your questions and can help you find an FH specialist if needed.

**General Treatments Goals for FH:**

For people with FH with no history of heart disease or stroke, the goal LDL-C is less than 100 mg/dL.

For people with FH who have heart disease or stroke, the goal is an LDL-C of less than 70 mg/dL. Recent recommendation for LDL-C is less than 55 mg/dL in higher risk people.

In general, the lower the LDL level the better.

**For family members:**

Because you have FH, your family members are also at risk of having FH. Close family members (parents, siblings, and children) all have a 50% chance of having FH and should consider screening. If your relatives need help finding screening, we are happy to help them with this.

**Staying Healthy**

In addition to maintaining an acceptable level of blood cholesterol through the use of cholesterol lowering medication

you can help lower your risk of heart disease and other conditions by following a healthy diet, engaging in exercise regularly, maintaining healthy levels of blood sugar and blood pressure, not smoking, and keeping your weight in a healthy range. Your health care provider can provide you with more guidance on these lifestyle measures.

**<<Resources** FamilyHeart.org>>

Sincerely

***

Contact Info

Genotype(-), Phenotype(+) Results Letter to Relative-

Dear _____,

Thank you for taking part in this Research Program involving Cascade Family Screening to Find

Familial Hypercholesterolemia (FH).

It was a pleasure working with you to help arrange your screening for FH. This letter is for your records, and it summarizes the results of your screening. A copy of your test results is included, and you are encouraged to share this with your health care provider.

**Summary:**

Based on your ***screening results, personal history, and family history, it is **probable** that you have familial hypercholesterolemia.It is important to understand even with a negative genetic test you can still be diagnosed with FH. It is important that you speak with your health care provider about these results. Early treatment with cholesterol lowering medications are available and they will help lower your risk for heart disease and stroke.

It is known that individuals with FH, if left untreated are at a greater risk for heart attack and stroke.

Lab Results:***- LDL above 190 mg/dL

Genetic testing- negative

Medications: ***

Personal History: ***

Family History: *** positive for early cardiac disease in first degree relative

**About Familial Hypercholesterolemia:**

FH can be diagnosed based on clinical findings therefore it should be treated even with a negative genetic test. Currently not all genes for FH have been identified.

Familial hypercholesterolemia (FH) is a common genetic condition that causes very high low density lipoprotein (LDL-C) cholesterol or “bad cholesterol”. FH is inherited, meaning it is passed down from parents to children.

**Next Steps:**

It is important to speak with your health care provider about these results.

There are a number of FH specialists who you might consider seeing as well.

An FH specialist will be able to work with you to determine the right treatment plan for you. The Research Team is here to answer your questions and can help you find an FH specialist if needed.

**General Treatments Goals for FH:**

For people with FH with no history of heart disease or stroke, the goal LDL-C is less than 100 mg/dL.

For people with FH who have heart disease or stroke, the goal is an LDL-C of less than 70 mg/dL. Recent recommendation for LDL-C is less than 55 mg/dL in higher risk people.

In general, the lower the LDL level the better.

**For family members:**

Because you likely have FH, your family members are also at risk of having FH. Close family members (parents, siblings, and children) all have a 50% chance of having FH and should consider screening that includes a lipid profile. If your relatives need help finding screening, we are happy to help them with this.

**Staying Healthy**

In addition to maintaining an acceptable level of blood cholesterol through the use of cholesterol lowering medication

you can help lower your risk of heart disease and other conditions by following a healthy diet, engaging in exercise regularly, maintaining healthy levels of blood sugar and blood pressure, not smoking, and keeping your weight in a healthy range. Your health care provider can provide you with more guidance on these lifestyle measures.

**<<Resources** FamilyHeart.org>>

Sincerely

***

Contact Info
