## Appendix 14 for "Patient and Clinician Perspectives on Centralized Cascade Screening for Familial Hypercholesterolemia in the United States: A Qualitative Implementation Study"

September 2, 2025

Dear Healthcare Professional,

**Your patient’s relative has been diagnosed with Familial Hypercholesterolemia (FH).**

**FH is a genetic condition** that causes very high lifelong LDL-cholesterol levels and is associated with increased risk for heart disease, heart attack, stroke, and premature death, if left untreated. First-degree relatives have a **50% chance** of having FH. More distant relatives are also at risk. If left untreated, people with FH have up to **20 times** the risk of developing premature heart disease. **Risk of premature death and cardiovascular events, such as heart attack and stroke, can be reduced by as much as 80% with early and aggressive lipid lowering therapy.^1^**

Your patient’s relative received a positive result for FH through genetic testing via **DISCOVER-FH**, a federally funded grant through University of Texas Southwestern (UTSW). As FH is an inherited condition, **we are offering your patient both a lipid panel and genetic testing at no cost.**

We are providing you with this information as your patient is likely to want to discuss this with you as their trusted source of healthcare information.

The following pages will give you more information about the DISCOVER-FH program at UTSW. Invitae, the MyCode program, and how to place an order for the genetic test. **Included with this information is your patient’s relative’s genetic testing lab report.**

We also strongly suggest that your patient has their **LDL-cholesterol checked, regardless of whether genetic testing is pursued.**

If you have any questions or concerns, please do not hesitate to contact us at **1 (844) 250-8031** or ****.

Sincerely,

Zahid Ahmad, MD

**Assistant Professor of Medicine**

**UTSW**

**Principal Investigator,** **DISCOVER-FH**

(xxx) xxx-xxxx |

^1^ Versmissen J, Oosterveer DM, Yazdanpanah M, et al. Efficacy of statins in familial hypercholesterolaemia: a long term cohort study. *BMJ*. 2008;337:a2423. Published 2008 Nov 11. doi:10.1136/bmj.a242

**Healthcare Professional FAQs**

**Can You Tell Me More About DISCOVER-FH?**

**DIrect SCreening Of RelatiVEs to Reveal FH (DISCOVER-FH)**

DISCOVER-FH is a federally funded four-year grant. The goal of DISCOVER-FH is to increase the number of at-risk individuals who are screened, diagnosed, and referred for treatment of FH. This program is modeled after a successful program in Holland and has been adapted for the United States. UTSW has partnered with the Family Heart Foundation, a national non-profit, research and advocacy organization, to contact at-risk relatives throughout the US and arrange for their genetic and lipid testing. The Family Heart Foundation will return test results to at-risk relatives and if positive will explain the importance of treatment. Dr. Zahid Ahmad from UTSW and Dr. Mary McGowan from the Family Heart Foundation are available to answer questions you may have about lipid lowering therapies and if needed can recommend a lipid specialist in your area for consultation.

**Can You Tell Me More About Genetic Testing Through This Program?**

**A genetic test is a clear way to determine if your patient has inherited FH.**

| 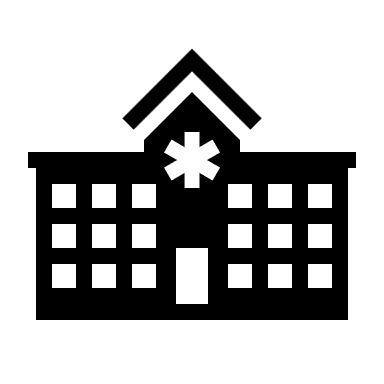 | Your patient’s relative received their FH result from UTSW. UTSW is a hospital system in Texas. UTSW has partnered with Labcorp, a nationwide laboratory, to provide genetic testing.     - Your patient is eligible for no cost genetic testing through Labcorp for a limited time. |
| --- | --- |
| 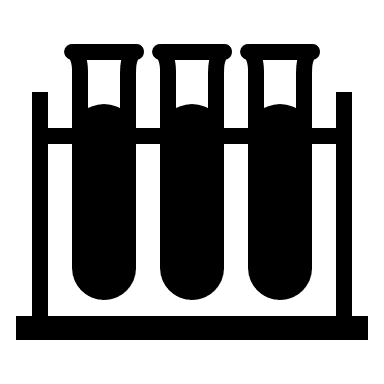 | The genetic test through Labcorp is performed on either saliva or blood.   - A member of the Family Heart Foundation will work with your patient to arrange for genetic testing. |
| 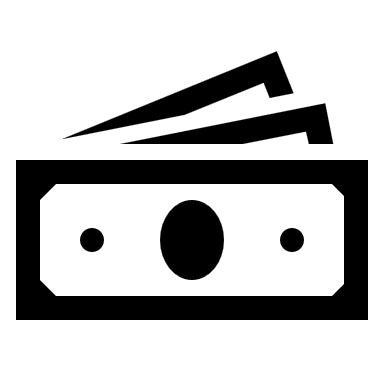 | The cost of your patient’s Family Variant genetic test is covered by Labcorp **if it is ordered by a member of the Family Heart Foundation team** {End Date of 150-Day Window}.   - Your patient and/or their insurance will be responsible for any potential costs associated with testing outside of this window or at a different laboratory. - Your patient and/or their insurance will be responsible for any potential costs associated with visiting a healthcare professional. |

**Does My Patient Also Need Cholesterol Testing?**

**Yes, they do! A cholesterol panel that includes LDL-C can also help you determine if your patient has FH.**

| 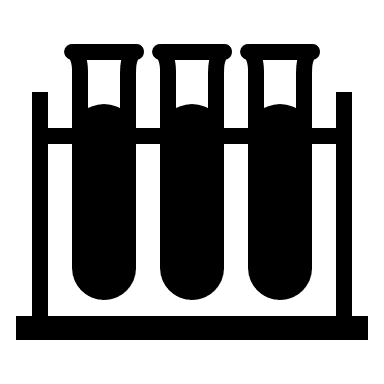 | Regardless of whether your patient gets a genetic test, we strongly suggest that they have their cholesterol checked. A lipid panel is also covered by Labcorp.   - A member of Family Heart Foundation can arrange for your patient to have a cholesterol panel that includes an LDL-C level drawn at a Labcorp facility. - A member of the Family Heart Foundation will call your patient to review the results of their cholesterol panel - If your patient is diagnosed with FH, cholesterol lowering medications will likely be required. |
| --- | --- |
| 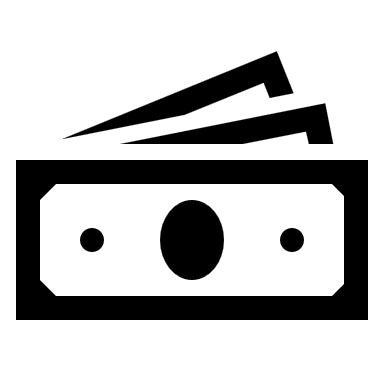 | Cholesterol testing and visits with a healthcare professional will not be covered by this program.   - Your patient and/or their insurance will be responsible for any potential costs associated with the cholesterol test or visits with a healthcare professional. |

**Healthcare Professional FAQs**

**How Can I Order A Genetic Test For My Patient?**

**The links below will give you step-by-step instructions on how to order your patient’s Family Variant genetic test through Invitae.**

Please use Code “**IMPACTFH**” when ordering.

For any additional questions and ordering assistance, please contact **Invitae’s client services**.

| 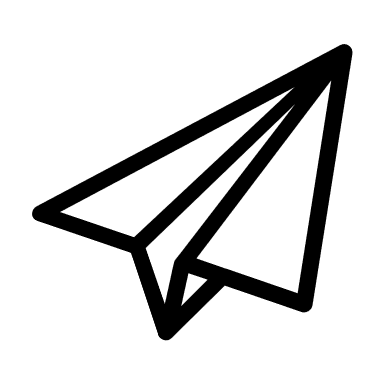 | ***Email***  | 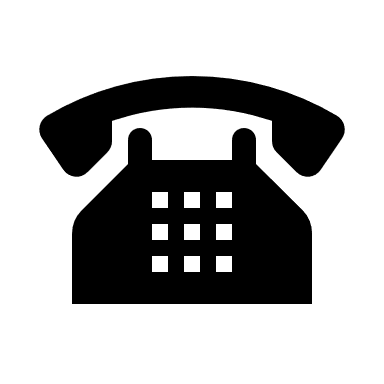 | ***Phone***  1 (800) 436-3037 |
| --- | --- | --- | --- |

| 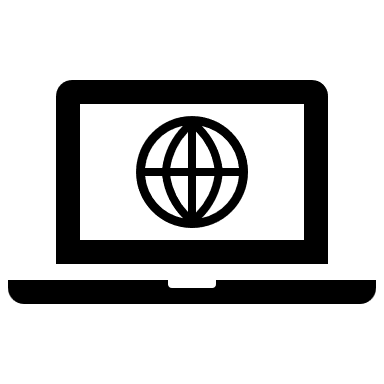 | ***Invitae’s Online Ordering Instructions*** invitae.com/en/ordering | ***Invitae’s Family Follow-up Testing Information***  invitae.com/en/family |
| --- | --- | --- |

**What If I Have Questions About FH Or This Program?**

**It is important to understand this information and why this is so important for your patient and their family. We are here to help!**

The **Geisinger Team** is here to help provide more information about FH and answer your questions about this program.

| 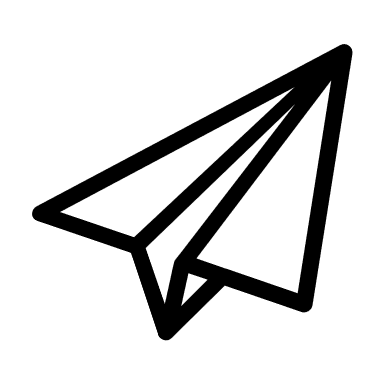 | ***Email***  | 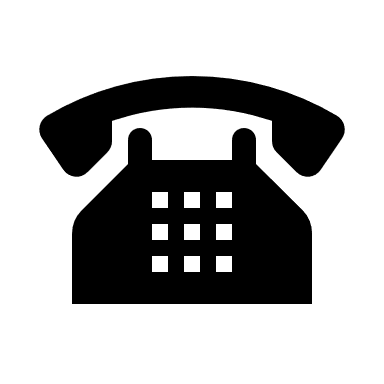 | ***Phone***  1 (844) 250-8031 |
| --- | --- | --- | --- |

| 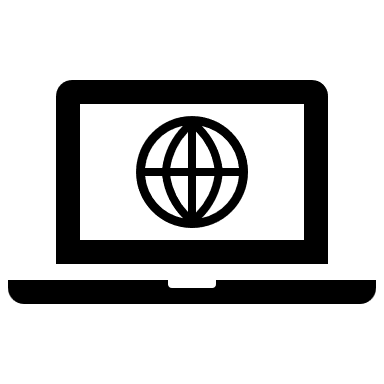 | ***Geisinger FH Webpage***  geisinger.org/FH |
| --- | --- |

The **FH Foundation** is a patient-centered organization that can help answer your questions about FH and provide support for you and your patient.

| 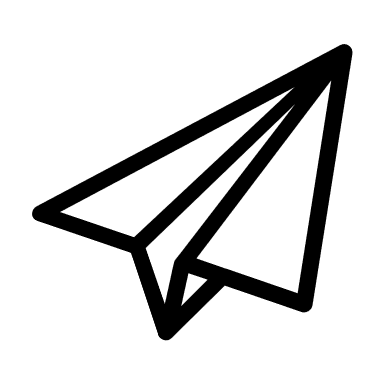 | ***Email***  | 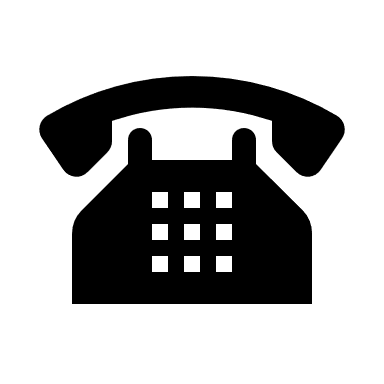 | ***Phone***  1 (626) 583-4674 | 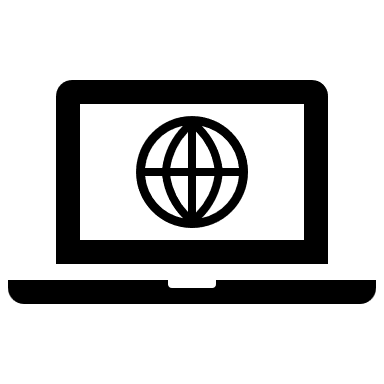 | ***Website***  theFHfoundation.org |
| --- | --- | --- | --- | --- | --- |

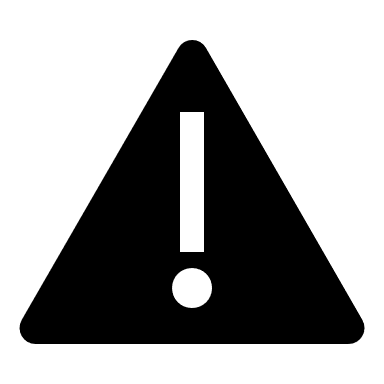

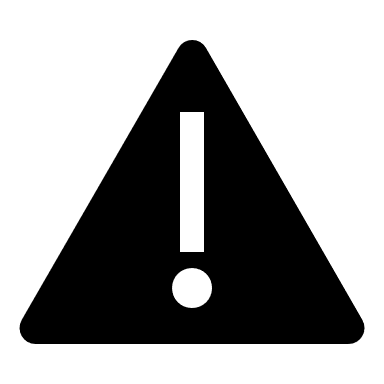

**The following page contains your patient’s relative’s Invitae lab report.**

**The information from the genetic test result will be needed to order your patient’s genetic test.**

**Please use Code “IMPACTFH” when ordering.**
