## Appendix 4 for "Patient and Clinician Perspectives on Centralized Cascade Screening for Familial Hypercholesterolemia in the United States: A Qualitative Implementation Study"

Pre-Test FH Counseling (Known FH Diagnosis for Discover FH)

**Inheritance:**

Heterozygous FH (HeFH) is inherited in an autosomal dominant manner. HeFH affects 1:250 individuals. Almost all affected individuals inherit the familial mutation from an affected parent. Children and siblings of an affected individual have a 50% chance of inheriting FH.

***Add in HoFH? Also, if we include the ABCG5/8 genes.

**Genetic Testing and Cardiovascular Disease Risk Management**

The choice to pursue genetic testing is always voluntary.

The following results are possible:

1. Positive result: A positive result (pathogenic or likely pathogenic variant) confirms the clinical diagnosis of familial hypercholesterolemia. Because there is a 50% chance that an individual’s parents, siblings, and children have also inherited this variant, genetic testing (for the specific variant detected) can be performed on these individuals to determine who else in the family is at risk.
2. Negative result: A negative result means no disease-causing variants were identified in the tested genes. A person clinically diagnosed with FH receiving a negative test result does not mean that they do not have FH—it simply means that their genetic cause(s) were not identified with current knowledge and current genetic testing technologies. About 30-40% of people with FH may test negative. At risk members should still have lipid screening and inform their doctors of FH in their family.
3. Variant of Uncertain Significance (VUS): Variants of uncertain significance are differences in a person’s DNA for which the clinical significance is unknown. There is not enough information at this time to say whether this difference causes FH or is a harmless, benign change. A VUS is not used to make medical management recommendations nor to offer genetic testing to family members. At risk members should still have lipid screening and inform their doctors of FH in their family.

**Limitations and risks:**

Genetic testing does not rule out all potential genetic causes for familial hypercholesterolemia. Our knowledge about the genetic causes for hypercholesterolemia continues to evolve. It is possible there are variants which have yet to be identified which may cause familial hypercholesterolemia. It is also possible that a person’s familial hypercholesterolemia is polygenic in nature, or, the result of many genetic changes each contributing only a small component for which testing is not yet available.

***Potential for non-paternity/revealing complex family dynamics??

**Genetic Information Non-discrimination Act:**

Provide information about GINA, the Genetic Information Non-discrimination Act, and its provisions against discrimination based on genetic status for employment and health insurance, and that it does not cover other sorts of insurance, such as life insurance and long-term disability insurance.

For active-duty service members: TRICARE may not use genetic information for coverage, underwriting, or premium-setting, but eligibility for TRICARE insurance is contingent upon employment by the U.S. Military, and GINA’s employment protections do not apply to the U.S. Military. The U.S. military is permitted to use genetic and medical information to make employment decisions.

***APOE **Secondary Findings:** It is possible that genetic testing for familial hypercholesterolemia will uncover unexpected results related to conditions outside of familial hypercholesterolemia. For example, certain variants in the *APOE* gene are also known to be a risk factor for Alzheimer’s disease. ***Sitosterolemia if including ABCG5/8.

**Lab: TBD*****

**Genes:** *LDLR*, *APOB*, *PCSK9*, *LDLRAP1*, *** *APOE*, *ABCG5*, *ABCG8*

**Cost:** Genetic testing through this research study, DISCOVER FH, is at no-cost to patients

**Logistics:** Blood draw/ saliva / buccal swab

We obtained the proper consent and the patient stated that they understood the information that was discussed and any questions were answered.
